# Frequency and neural correlates related to psychosis in motor neurone disease

**DOI:** 10.1101/2021.03.31.21253701

**Authors:** A. Wilcox, P. S Jones, R. C. Roberts, J. B. Rowe

## Abstract

Psychosis is a challenging feature of the syndromes motor neurone disease (MND), frontotemporal dementia and their overlap. Clinically evident psychosis affects 5-10% of patients, and more in those with C9*orf*72 expansions. However, subthreshold psychosis features may be overlooked in the context of overriding concern for physical impairment. This prospective study aimed to establish the prevalence and severity of psychosis features in a population-representative sample of MND, and to identify the neural correlates of psychosis by structural magnetic resonance imaging. A three-tiered system was applied to recruit people with MND, with cognitive and psychosis screening (Tier 1: *N*=111 with the Edinburgh ALS Cognitive and Behavioural Screen), in-depth neuropsychiatric assessment (Tier 2: *N*_-_=60) and imaging (Tier 3: *N*=30). Age-, education- and sex-matched healthy controls were recruited to Tier 2 (N=30) and Tier 3 (N=20). Overt psychosis was identified in 10% of the Tier 1 cohort, whilst 46% showed milder and diverse neuropsychiatric change. Grey matter correlates of psychosis included atrophy of the cingulate cortex and the hippocampus. White matter correlates included compromised integrity along frontotemporal and temporal-parietal association pathways, especially those connecting the anterior temporal lobe. These grey and white matter changes in MND represent vulnerability to psychosis and are qualitatively similar to volumetric and white matter abnormalities observed in other primary psychotic disorders. Neuropsychiatric features are common, even though overt psychosis is identified in a minority of people with motor neurone disease.

## INTRODUCTION

Psychosis is a challenging clinical aspect of motor neurone disease (MND), frontotemporal dementia (FTD) and their overlap (FTD-MND). Psychosis is more commonly described in FTD than MND, with higher prevalence associated with C9*orf*72 hexanucleotide expansions across the continuum of FTD-MND (Devenney et al., 2017; Snowden et al., 2013). The focus on familial MND from C9*orf*72 mutations reflects the high prevalence of psychosis, in up to a third of patients. However, the prevalence of psychosis and related neuropsychiatric features in sporadic MND is not well established.

Psychosis associated with MND may lead to an initial diagnosis of a primary psychotic disorder such as schizophrenia, or somatoform psychosis (Arighi et al., 2012; Velakoulis et al., 2009). Phenomenologically, it is similar to psychosis observed in FTD and FTD-MND, with delusions (persecutory and somatic delusions, pathological jealousy and grandiosity) and hallucinations (second person auditory, visual hallucinations of people and animals; tactile hallucinations). However, more commonly reported are milder, subthreshold psychosis features with suspiciousness, somatisations (e.g. “crawling” sensation on or beneath the skin), fixed or overvalued ideas about comorbid illness (Devenney et al., 2014; Snowden et al., 2012). These latter symptoms are subtler but more common than in primary psychotic disorders.

The neurobiology of psychosis in MND is not well characterised, but insight can be gained from other psychotic conditions, such as schizophrenia. Chronic schizophrenia is associated with widespread atrophy in frontal, temporal, limbic, striatal and cerebellar structures (Shenton et al., 2001). These areas may be abnormal even at the early emergence of symptoms in people with a first episode psychosis (Meisenzahl et al., 2008; Torres et al., 2016). Both positive and negative symptoms have been correlated with regional atrophy (Modinos et al., 2013) and with widespread abnormalities in diffusion properties in cerebral white matter tracts connecting the frontal and temporal lobes (Skelly et al., 2008). Auditory hallucinations have consistently been related to volume reduction of the medial and superior temporal lobe and adjacent insula (Modinos et al., 2013; Van Tol et al., 2014), and delusions related to abnormal diffusion in the uncinate fasciculus (Nakamura et al., 2005).

To identify the neural correlated of psychosis in MND, this study tested *a priori* regions of interest and their connections in adjacent white matter. The regions of interest were selected as candidate mediators of three pro-psychotic processes: (i) the amygdala, as part of the limbic system associated with aberrant associative learning and attribution of causality; (ii), orbitofrontal cortex, nucleus accumbens and the caudate, as part of a frontostriatal circuit mediating valence and attentional control; and (iii) the cerebellum, associated with sensorimotor integration and somatisation.

Previous imaging studies of psychosis or related neuropsychiatric behaviour in FTD-MND have stratified patient by diagnosis (Devenney et al., 2017), or by gene status such as C9*orf*72, *MAPT*, or *GRN* (Sellami et al., 2018). The focus in these studies has been on FTD, FTD-MND and/or those with C9*orf*72 mutations, in which psychosis is more commonly reported (Devenney et al., 2017; Snowden et al., 2012). Here we focus instead on MND, without genotypic selection. In a regional specialist service for people affected by MND, we undertook cognitive and psychosis screening (Tier 1), then in-depth neuropsychiatric assessment (Tier 2) and structural brain imaging (Tier 3).

## METHODS

### Participants

Participants were recruited through a tiered system, shown in Figure 1, embedded in a regional healthcare service (*N*=186 during the study period), where we introduced research screening for cognition change and psychosis (*N*=111), followed by neuropsychological and structured neuropsychiatric assessment (*N*=60) and structural brain imaging (*N*=30). 30 age-, sex-, education-matched healthy controls were also recruited.

**Figure 1:**
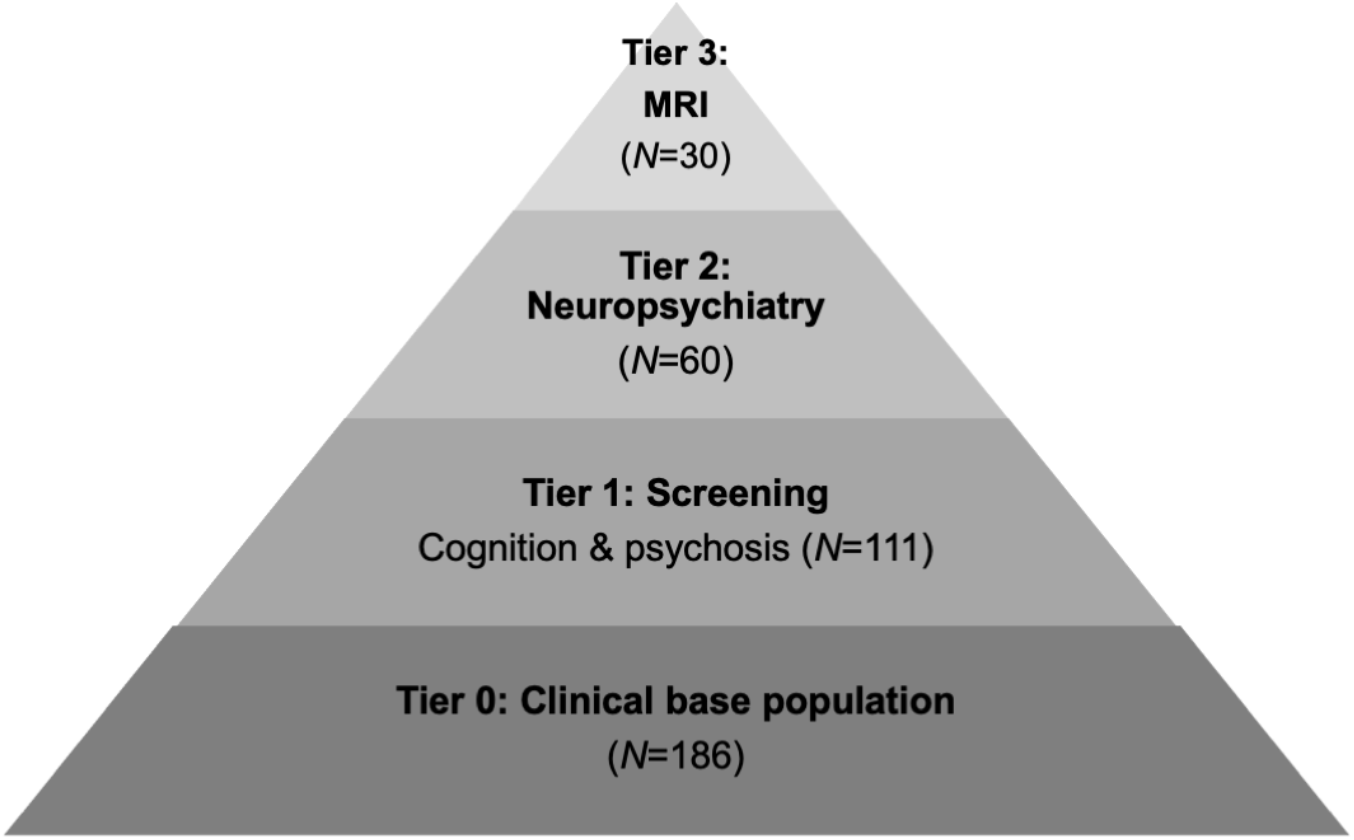
Overview of the study tiers.

Patients were assessed in a Clinical Research Facility or in their homes. Diagnosis of MND was made by the neurologist-led multidisciplinary team at the Cambridge MND Care Centre according to standard criteria (Brooks, 1994). Disease severity was assessed with the ALS-Functional Rating Scale-Revised (ALS-FRS-R) (Cedarbaum et al., 1999) and FTD-Functional Rating Scale (FTD-FRS) (Mioshi et al., 2010). Disease duration was estimated in months from symptom onset to cognitive screen date. Exclusion criteria included concomitant significant neurological or psychotic condition unrelated to MND, use of psychotropic medications, and previous history of drug or alcohol abuse. None of the patients in the study were taking anti-psychotic medication or psychosis-inducing medication at the time of their initial screen or testing session.

### Cognition, psychosis and neuropsychiatric measures

Participants underwent a standardised assessment battery, including the Edinburgh Cognitive and Behavioural ALS Screen (ECAS) (Abrahams et al., 2014a); the Brief Psychiatric Rating Scale (BPRS); Neuropsychiatric Inventory (NPI) (Cummings et al., 1994); and the Revised Cambridge Behavioural Inventory 36-item version (CBI-R) (Wear et al., 2008). Two psychosis index scores were derived. One, by combining the *Delusions* and *Hallucinations* sub-scores on the NPI and the second, by combining the sub-sections, *Abnormal Behaviour, Beliefs*, and *Stereotypical and Motor Behaviours* on the CBI-R. Mood was assessed using the Beck 21-item Depression Inventory (BDI-II) (Beck et al., 1961) and the Hospital Anxiety and Depression Scale (HADS) (Zigmond & Snaith, 1983). Apathy was measured using a recently developed Cambridge Questionnaire for Apathy and Impulsivity (Cam-QuAIT) and the Dimensional Apathy Scale (DAS) (Radakovic & Abrahams, 2014).

### Neuroimage acquisition and pre-processing

Tier 3 participants (*N*=30 patients, *N*=20 controls), underwent whole-brain structural imaging at the Wolfson Brain Imaging Centre, University of Cambridge, UK using a Siemens 3T PRISMA scanner (Siemens Healthcare, Erlangen, Germany) with a 64-channel head coil. T1-weighted MPRAGE images were acquired with repetition time (TR) = 2000ms, echo time (TE) = 2.93ms, acquisition matrix of 256×256, 208 slices of 1.1mm thickness, voxel size 1.102×1.102×1.1mm^3^, inversion time = 850ms and flip angle = 8°. T2-weighted images were acquired with TR = 3200ms, TE = 401ms, acquisition matrix of 256×256, 176 slices of 1.1mm thickness, voxel size 1.102×1.102×1.1mm^3^, and flip angle = 120°. Two repeats of 64 volumes of diffusion weighted gradient images were acquired in 64 directions with *b* value 1000s/mm^2^ and five volumes without diffusion weighting, TR = 7300ms, TE = 90ms, axial in-plane acquisition matrix of 96×96, 59 slices of 2.5mm slice thickness (no gap), voxel size 2.5×2.5×2.5mm^3^, flip angle = 90°.

Grey matter volume was studied from cortical parcellation and subcortical segmentation in FreeSurfer version 6.0.0 (http://surfer.nmr.mgh.harvard.edu/), using the automated *recon-all* script, applying the cortical labels from the Desikan-Killiany Cortical Atlas. Processing included motion correction and averaging of T1 and T2-weighted images, normalisation, skull stripping, and removal of non-brain structures, followed by the construction of models of the boundary between white matter and cortical grey matter and the pial surface (grey matter to CSF). T2-weighted images were used in combination with T1 to increase pial definition of cortical areas. Diffusion weighted image processing used FMRIB Software Library (FSL) diffusion toolbox (version 5.0.9; www.fmrib.ox.ac.uk/fsl), to study Mean Diffusivity (MD) and Fractional Anisotropy (FA). A brain mask was created from first B0 image of the DWI series and all volumes were adjusted for eddy currents and head movements using *eddy*. Then diffusion maps were created by voxelwise fitting of the diffusion tensor using FSL *dtifit*.

### Statistical analyses

Statistical analyses of group difference and correlations used R Studio version 3.6.1 (2019-07-05). Group differences between MND and healthy controls on clinical characteristics, demographics, neuropsychiatric, grey matter volumes, including investigations of the differential relationship of psychiatric and cognitive outcome measures on regional brain volume used linear mixed models. Analyses of white matter diffusion metrics used Tract-based spatial statistics (TBSS) in the FSL diffusion toolbox with FSL *randomise*. Covariates of no interest included age, total intracranial volume and disease severity.

False Discovery Rate (FDR) correction and Threshold-Free Cluster Enhancement (TFCE) with 2D optimisation were applied for multiple comparisons of FreeSurfer output and TBSS respectively. Assumptions were tested using Shapiro Wilk test of normality and inspecting histograms/boxplots and Q-Q Normality Plots, together with Bartlett’s test of homogeneity of variance (where outcome measures were non-normal, Levene’s test of homogeneity of variance was applied). Parametric tests with grey matter volumes used arcsine transformation. Non-parametric Wilcoxon test and Spearman’s *rho* correlations were applied to CBI-R and NPI total and psychosis index scores due to non-gaussian distributions.

## RESULTS

Table 1 describes the participants across each tier, for age, education, disease duration, severity, survival and cognition, sex, MND sub-type, and presence of psychosis. In Tier 2, people with MND and healthy controls were matched for age and education, but with a mild excess of male MND participants (Table 3). In Tier 3 participants were matched by age and sex. Sex and age were included as covariates in subsequent analyses to account for any variance in these demographics. People with MND demonstrated the expected range of functional and cognitive impairment, denoted by reduced ALS-FRS-R, FTD-FRS and total ECAS scores, compared to controls.

**Table 1:**
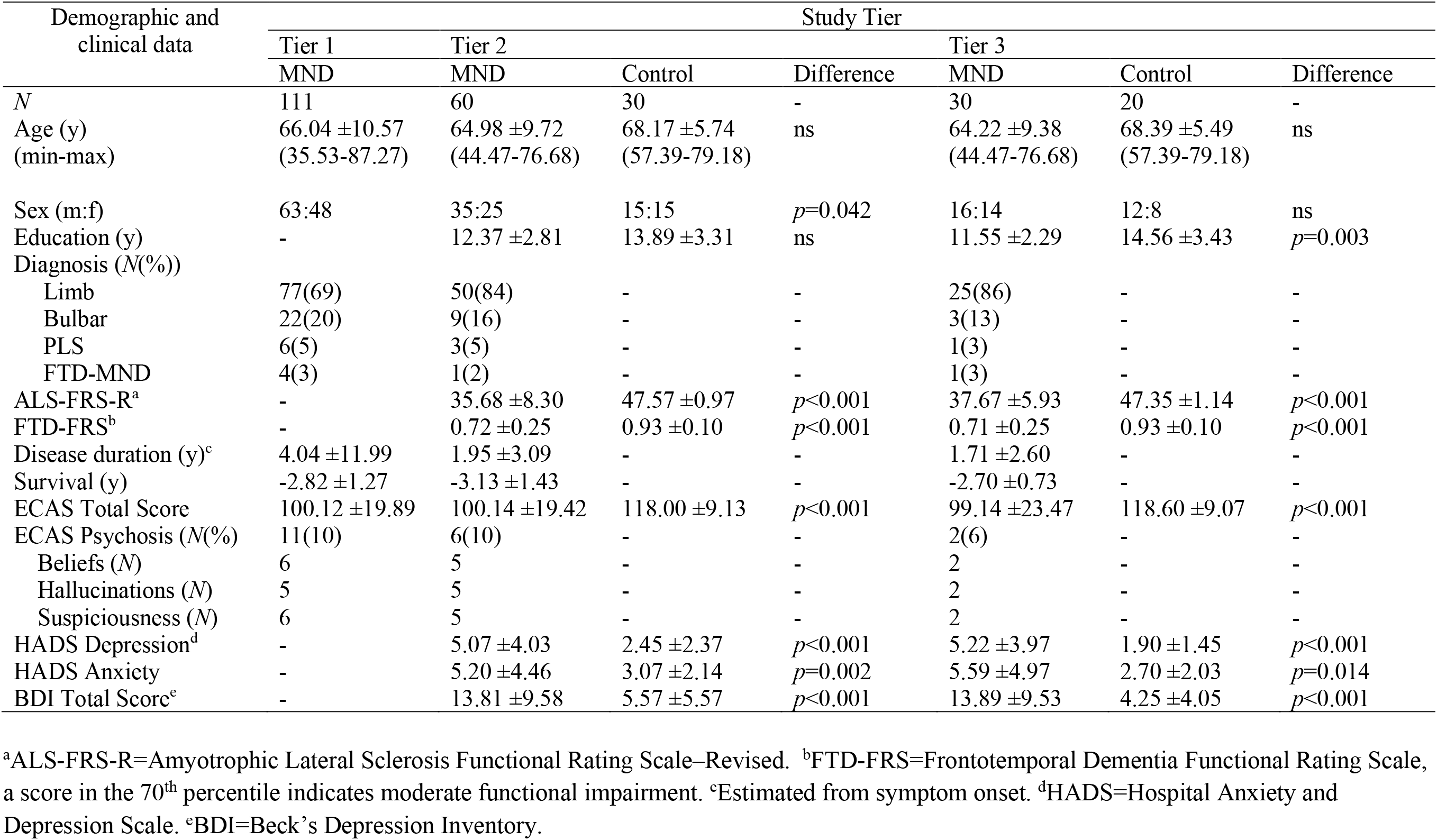
Clinical characteristics and group differences across Tiers 1-3.

### Psychosis and neuropsychiatric group comparisons

The prevalence and details of the neuropsychiatric features in each Tier are shown in Table 1. The carer report on the ECAS cognitive screen indicated 10% of people with MND in Tier 1 endorsed at least one overt psychosis feature (e.g. “has strange and/or bizarre beliefs and behaviours”, “hears or sees things that are not there, and/or feels the presence of someone who is not there”, and “Is overly suspicious, and/or feels persecuted”). The prevalence of psychosis was similar in Tier 2, but lower for Tier 3. Figure 2 presents the behavioural items from the ECAS for Tiers 1-3 in people with MND. Behavioural change, as observed by someone who knows the patient well, was noted in half of the MND group. Apathy was the most common behavioural change, affecting a quarter of people with MND. Additionally, people with MND showed other odd and bizarre behaviours, such as social inappropriateness, perseverate, repetitive movements and compulsive behaviours.

**Figure 2:**
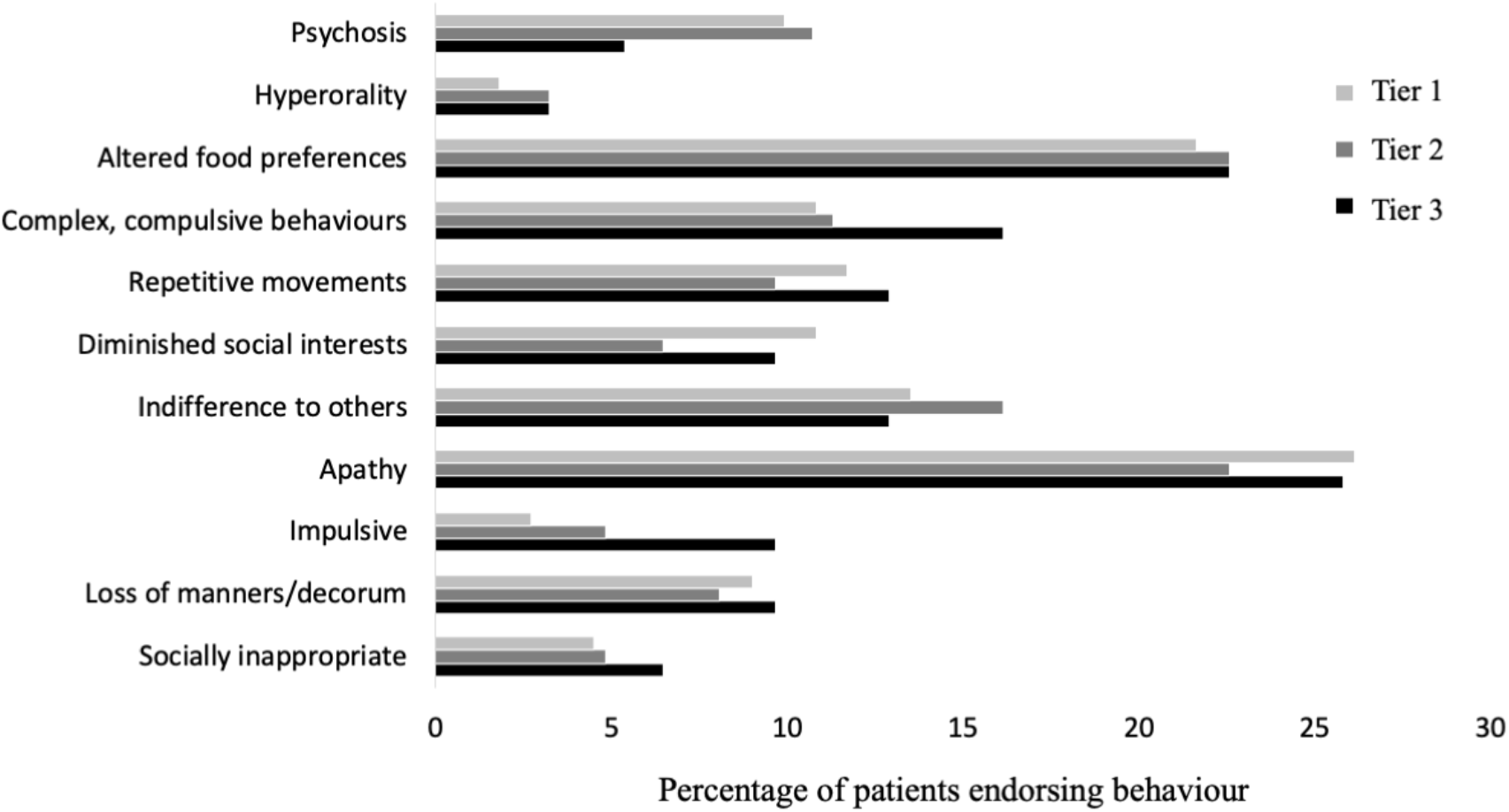
Percentage of Tier 1, 2 and 3 people with MND for whom each behaviour was endorsed by a carer.

Group differences were confirmed in the psychosis scales for Tier 2 participants (CBI-R, NPI and BPRS, Table 1). Figure 3 illustrates the variation in scores from each person with MND and control participant in Tier 2, overlaid onto group average box plots. This identifies 12-16% of MND participants to have mild to moderate psychosis.

**Figure 3:**
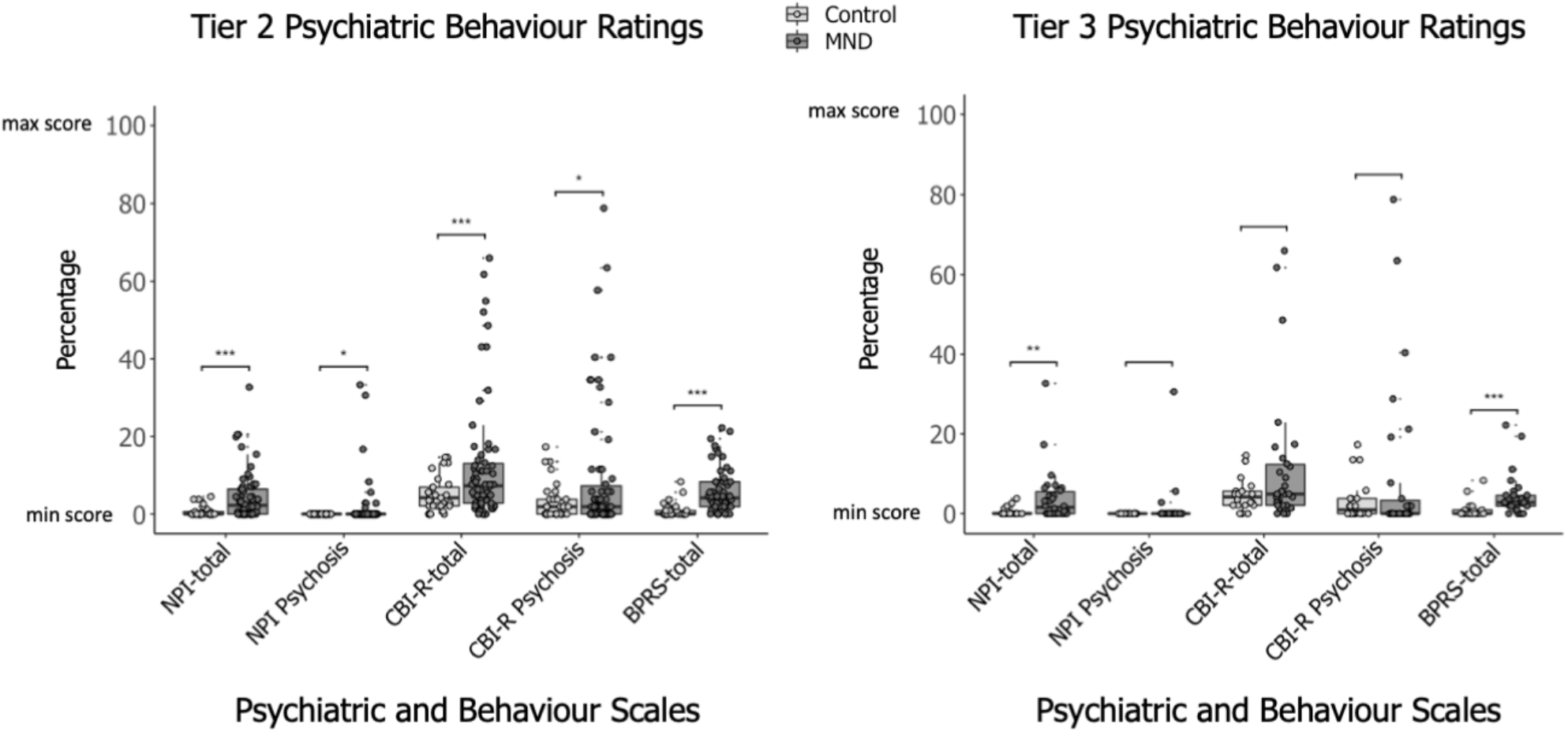
Tier 2 (left) and Tier 3 (right) variation in percentage of psychiatric and behaviour ratings across three scales for MND and control participants (scatter plot) overlaid onto group average ratings (boxplot). Ratings converted to percentage of total score of respective scales and rescaled from minimum 0 to maximum 100. Ratings closer to 100 indicate a higher involvement of psychiatric behaviour. NPI-total=Neuropsychiatric Inventory (total score), NPI Psychosis=NPI psychosis index, CBI-R-total=Cambridge Behavioural Inventory-R (total score), CBI-R Psychosis=CBI-R psychosis index, BPRS-total=Brief Psychiatric Rating Scale (total score). \**p*<0.05, \*\*\**p*<0.001.

### Neuroimaging

#### Grey matter volume analyses

Group comparison between people with MND and controls revealed grey matter volume loss by MND in previously reported extra-motor regions, including the *a priori* regions of caudate, amygdala, but not the orbitofrontal cortex (Table 2). Other regional differences were significant only with the uncorrected threshold.

**Table 2:** Averaged regional mean volumes (ml) by group.

| Region | MND<br>(N=30) | Control<br>(N=20) | T statistic | R<sup>2</sup> | Group<br>difference |
| --- | --- | --- | --- | --- | --- |
| Frontal Lobe | 152.80 | 159.97 | $t=-2.10$ | 0.42 | $p=0.042$ |
| Temporal Lobe | 102.01 | 109.26 | $t=-2.11$ | 0.54 | $p=0.031$ |
| Occipital Lobe | 46.16 | 49.72 | $t=-2.17$ | 0.58 | $p=0.035$ |
| Parietal Lobe | 104.37 | 107.53 | $t=-1.18$ | 0.60 | ns |
| Cingulate | 15.74 | 16.94 | $t=-1.27$ | 0.54 | ns |
| Orbitofrontal* | 22.88 | 24.22 | $t=-1.91$ | 0.56 | ns |
| Hippocampus | 7.05 | 8.63 | $t=-2.78$ | 0.50 | $p=0.007$ |
| Amygdala* | 2.89 | 3.26 | $t=-2.86$ | 0.49 | $p=0.037$ |
| Insula | 12.02 | 13.10 | $t=-1.56$ | 0.44 | ns |
| Caudate* | 6.44 | 7.15 | $t=-2.57$ | 0.48 | $p=0.041$ |
| Nucleus Accumbens* | 0.91 | 1.01 | $t=-1.54$ | 0.35 | ns |
| Thalamus | 12.98 | 13.15 | $t=0.11$ | 0.55 | ns |
| Putamen | 5.85 | 9.05 | $t=-1.09$ | 0.43 | ns |
| Cerebellum* | 100.46 | 105.11 | $t=-1.31$ | 0.68 | ns |

\**a priori* regions-of-interest based on hypotheses set out in introduction. Unpaired *t*-tests, uncorrected *p*-values. Differences did not survive FDR correction.

#### Grey matter correlates of psychosis

The degree of psychosis in MND was associated with reduced volume in two regions. There was an interaction between group and the CBI-R psychosis index on cingulate volume (*F*(1,44)=6.74, *p*=0.013) and hippocampus volume (*F*(1,44)=1.06, *p*=0.002) (Figure 4). No significant grey matter correlates were identified for the regions-of-interest.

**Figure 4:**
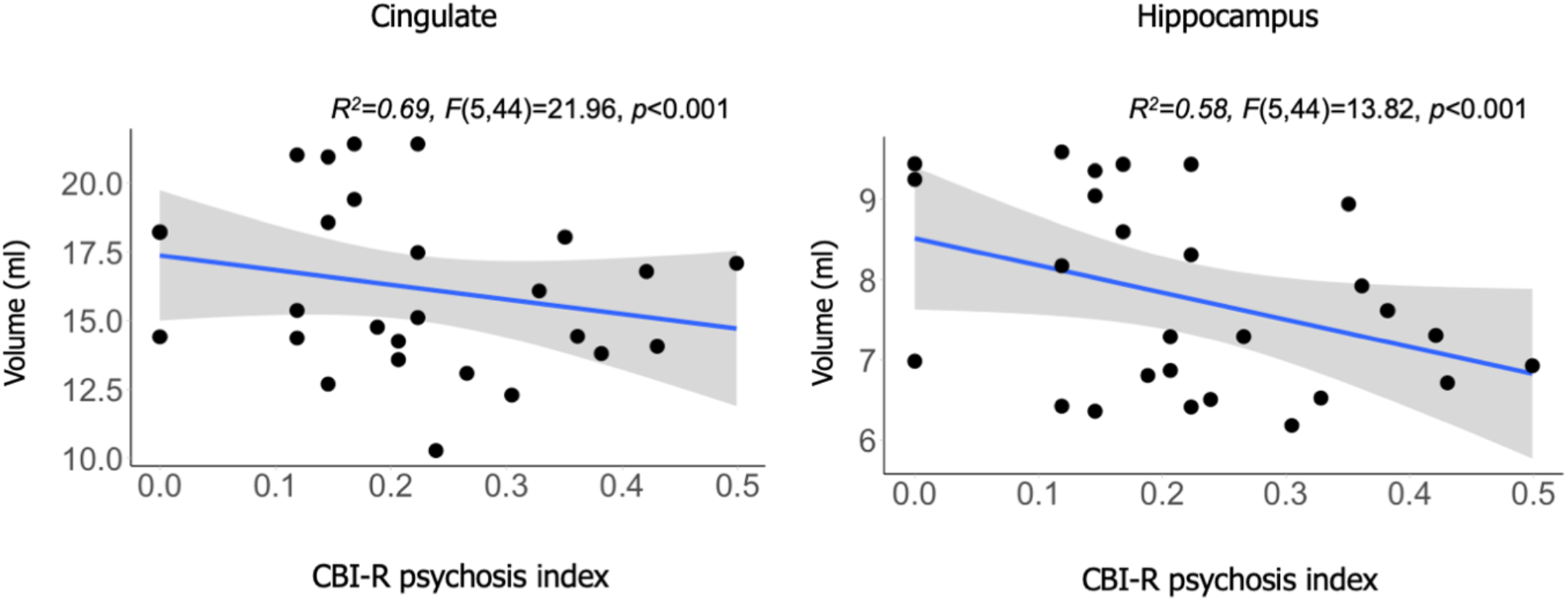
Within-group MND correlation between score on CBI-R psychosis index (arcsine transformed) on regional volume of cingulate (left) and hippocampus (right). R^2^=adjusted R^2^ for age and TIV. *F*-statistic=overall model. *p*-value unc.

#### White matter correlates of psychosis

Mean Diffusivity correlated with CBI-R psychosis index in the MND group in three clusters between 41 and 2,551 voxels (TFCE *p*<0.05). These clusters included peaks around the inferior longitudinal fasciculi and uncinate fasciculi of the bilateral anterior temporal cortex and temporal pole (cluster 1: 2,551 voxels, *t*=7.29, TFCE *p*<0.05; cluster 2: 114 voxels, *t*=9.56, TFCE *p*<0.05; cluster 3: 41 voxels, *t*=6.98, TFCE *p*<0.05; Figure 5). No interactions with the control group were identified. No significant decreases in Fractional Anisotropy were identified. There were no significant clusters associated with the NPI for Fractional Anisotropy or Mean Diffusivity.

**Figure 5:**
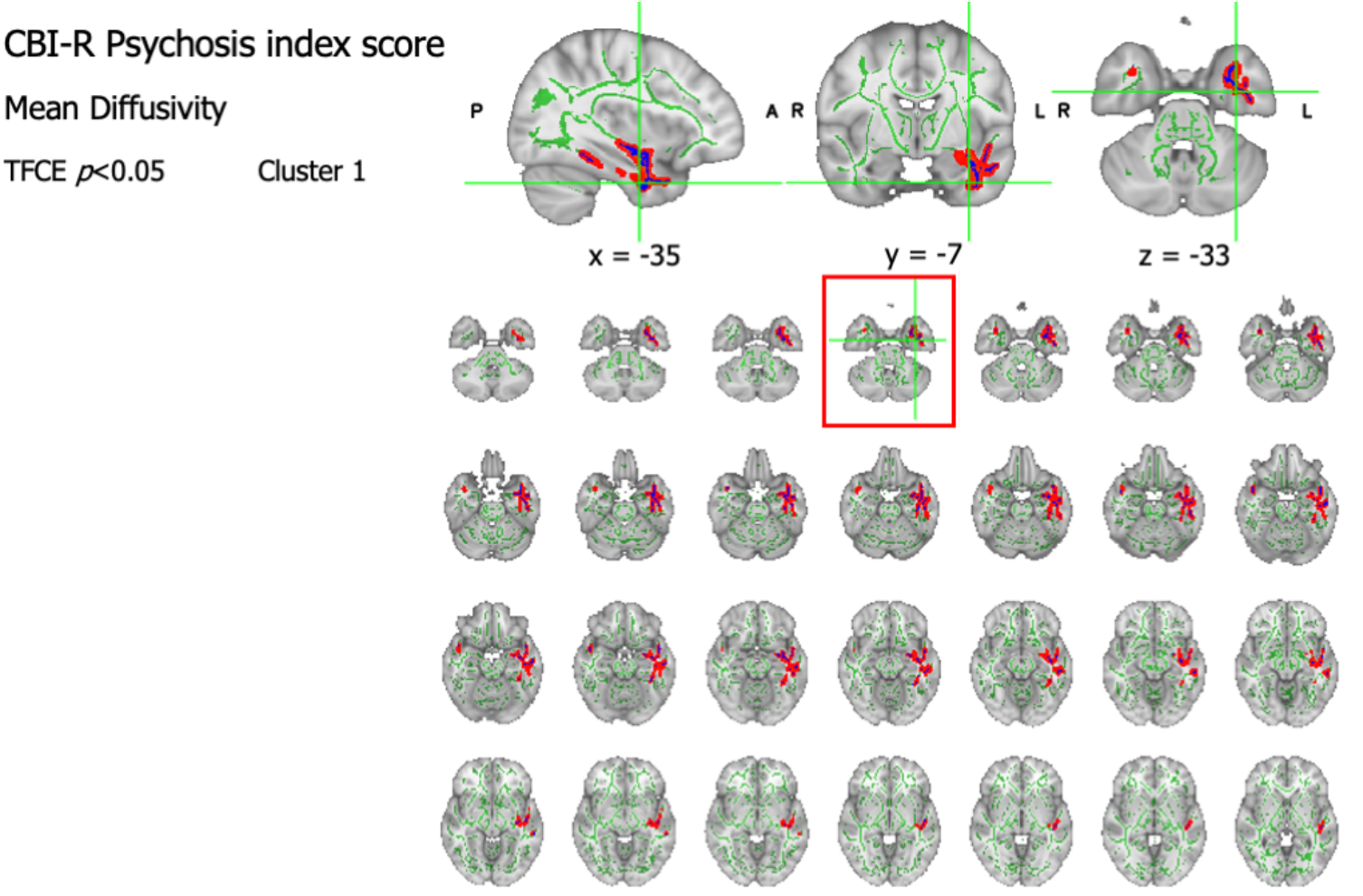
White matter correlates of psychosis in MND, with increased Mean Diffusivity (“fattened” skeletonised tracts in red, TFCE *p*<0.05). Crosshairs indicate highest intensity voxel and anatomical location given in MNI coordinates. Results overlaid on the mean FA skeleton (green) and MNI152 T1 template (grey). P=posterior, A=anterior, R=right, L=left.

## DISCUSSION

This study confirms a prevalence of psychosis of approximately 10% in people with MND; while overt presentations of psychosis may be uncommon in MND patients, a 10% prevalence suggests that screening is warranted. In addition, we found psychosis was related to regional changes in white matter diffusivity in the temporal pole, bilaterally and to atrophy of the cingulate and hippocampus. These complement the neural correlates of psychosis in FTD and FTD-MND (Devenney et al., 2017; Sellami et al., 2018) and accord with structural associations of other psychotic conditions.

The screening program identified psychosis in approximately 10% of MND based on the ECAS assessment, which refer to visual/auditory hallucinations, bizarre beliefs or behaviours and persecutory ideation. This 10% rate is higher than previous reports by Lillo et al., (2011) (5%), Crockford et al., (2018) (2.9%), and Abrahams et al., (2014b) and McHutchison et al., (2019) (both *N*=1). Previous studies of the continuum of FTD through to MND have focussed on sub-groups where psychosis is more prominent. Psychosis more commonly occurs in FTD and FTD-MND, with prevalence of 30-60% associated with C9*orf*72 gene expansion, (Devenney et al., 2017; Snowden et al., 2013). The current rate of MND-psychosis (∼10%) is consistent with some studies based on FTD-MND (Sha et al., 2012), but other FTD-MND cohorts showed higher rates of 14 – 38% (Devenney et al., 2017, 2019; Saxon et al., 2017; Snowden et al., 2012), which may have been driven by those with overlapping FTD symptoms. Some FTD-MND studies report a lower incidence of ∼5% (Mahoney et al., 2012). This study confirms that even in the absence of meeting diagnostic criteria for concomitant FTD, patients with MND may be psychotic (Lillo et al., 2011).

Given that most people with MND do not have psychosis, we examined the neural correlates of the severity of psychosis or pro-psychotic symptoms. Although our *a priori* regions of interest caudate and amygdala were atrophic in MND at the group level, their volume did not correlate with psychosis. Instead, grey matter atrophy correlation was identified in the hippocampus and cingulate, at least in the exploratory analyses. In MND, the hippocampus and cingulate have consistently been implicated in other neurobehavioral disorders, by neuroimaging (Bede et al., 2013; Kato et al., 1993), neuropsychology (Consonni et al., 2019; Machts et al., 2015; Phukan et al., 2012) and neuropathological studies (Brettschneider et al., 2012; Takeda et al., 2009). Further along the FTD-MND continuum, atrophy in these regions have been related to psychosis in FTD, and C9*orf*72-associated FTD-MND (Devenney et al., 2017) as well as broader changes in personality and behavioural changes, such as apathy and disinhibition with FTD (Consonni et al., 2019). The current results are consistent with and extend previous work on the investigation of psychosis in the continuum of FTD-MND.

The hippocampus and cingulate are of particular interest as these regions are vulnerable in prodromal psychosis in other conditions, and in the acute transition to overt psychosis (Pantelis et al., 2003; Smieskova et al., 2010). Neuroanatomical abnormalities in the medial temporal lobe and cingulate that predate the first episode of overt psychosis have been identified in at-risk individuals with prodromal symptoms (Pantelis et al., 2003).

Longitudinally, changes in these regions in at-risk individuals who did not subsequently develop psychosis have not been identified (Pantelis et al., 2003), indicating the medial temporal lobe and cingulate have a strong association with the development of psychosis. Additionally, these regions may play a role in the maintenance of psychotic symptoms as patients with established schizophrenia show qualitatively similar volume abnormalities in the hippocampus and cingulate (Torres et al., 2016).

The white matter of the temporal pole, uncincate fasciculus the inferior longitudinal fasciculus connect the amygdala and hippocampus to the frontal lobe, and to sensory perceptual systems (Catani & Thiebaut de Schotten, 2008). The white matter tract abnormalities associated with the features of psychosis identified in the current study are consistent with the grey matter correlates. They are also consistent with changes in the inferior longitudinal and uncincate fasciculi in early psychosis and at-risk individuals (Carletti et al., 2012; Cooper et al., 2018; Luck et al., 2011) and in schizophrenia (Phillips et al., 2009; Seitz et al., 2016).

The compromise of these white matter tracts in our study mirrors the correlations with positive and negative symptom severity in at-risk psychosis groups and primary psychotic conditions. For example, reduced Fractional Anisotropy in the uncinate fasciculus with positive (Bopp et al., 2017; Nakamura et al., 2005) and negative (Luck et al., 2011) symptom severity, and in the inferior longitudinal fasciculus with disordered thought (Phillips et al., 2009). The neural mechanisms involved in distinct psychosis symptoms in MND may differ and future research should further delineate psychosis features and their separate neural correlates.

There are limitations to our study. First, we only recruited two thirds of the patients seen in the regional MND Care Centre. Not everyone in the healthcare service was able or willing to participate in research, and this may introduce selection bias to our cohort. Second, our cohort was not large enough to reliably break down the psychosis prevalence by MND subtype, nor did we track individuals’ symptomatology longitudinally. Both would be valuable extensions in future work. Third, selection biases into Tiers 2 and 3 are suggested by the mild mismatching of sex or education. Although we adjusted for these confounds in our analysis, residual effects are possible. Fourth, none of our participants were taking antipsychotic medication. This suggests that their psychosis may not have been recognised, or was sufficiently mild in the context of other deficits, as to not require medication. This assumption may not be justified, as even in the absence of challenging behaviours arising from psychosis there may be distress or impacts on autonomous decision-making and care planning. Finally, the modest sample size in Tier 3 (N=30 for MRI) means that subtler structural consequences of MND and correlates of psychosis may not have been detected due to type II error.

In conclusion, psychosis affects approximately 10% of people with MND but individual variability is high. In clinical settings, both overt and milder psychosis symptoms may be overlooked, especially in the context of severe physical impairment. Yet psychosis can affect patients’ wellbeing, their psychological response to their condition, their decisional autonomy and carer stress. Our imaging results suggest that psychosis is related to multi-focal atrophy and compromised integrity of white matter association tracts centred on the anterior temporal lobe and its connections to the frontal cortex, in a network associated with other psychotic disorders. These homologies may be exploited to improve diagnosis and treatment options of psychosis in people with motor neuron disease.

## Data Availability

The datasets generated during and/or analysed during the current study are available from the corresponding author on reasonable request.

## ACKNOWLEDGMENTS

We would like to thank the following: all the people with MND, their families and carers and healthy control participants. The Cambridge MND care centre coordinating team, Dr Dirk Baumer and Dr Jonathan Tay.

## FUNDING

This research was funded by Alzheimer’s Research UK Studentship (PhD2017-26); Medical Research Council (SUAG 051/ G101400); and the NIHR Cambridge Biomedical Research Centre (BRC-1215-20014). The views expressed are those of the authors and not necessarily those of the NIHR or the Department of Health and Social Care.

## Notes

### Competing Interest Statement

The authors have declared no competing interest.

### Funding Statement

This research was funded by Alzheimers Research UK Studentship (PhD2017-26). Medical Research Council (SUAG 051/G101400). The NIHR Cambridge Biomedical Research Centre (BRC-1215-20014). The views expressed are those of the authors and not necessarily those of the NIHR or the Department of Health and Social Care.

### Author Declarations

London Queens Square Research Ethics Committee. Reference number: 14/LO/2045.

